# Beneficial Effects of novel Aureobasidium Pullulans strains produced Beta-1,3-1,6 Glucans on Interleukin-6 and D-Dimer levels in COVID-19 patients; results of a randomized multiple-arm pilot clinical study

**DOI:** 10.1101/2021.08.09.21261738

**Authors:** Kadalraja Raghavan, Vidyasagar Devaprasad Dedeepiya, Vaddi Suryaprakash, Kosagi-Sharaf Rao, Nobunao Ikewaki, Tohru Sonoda, Gary A. Levy, Masaru Iwasaki, Rajappa Senthilkumar, Senthilkumar Preethy, Samuel JK Abraham

## Abstract

**Objective:** Cytokine storm and Coagulopathy have been implicated as major causes of morbidity and mortality in COVID-19 patients. A black yeast Aureobasidium pullulans AFO-202 strain produced beta 1,3-1,6 glucan has been reported to offer potential immune enhancement and metabolism balancing, as well as mitigation of coagulopathy risks. The N-163 strain produced beta glucan is an efficient anti-inflammatory immune modulator. In this pilot clinical study, we report the beneficial effects of these two beta glucans on the biomarkers for cytokine storm and coagulopathy in COVID-19 patients.

**Methods:** A total of 24 RT-PCR positive COVID-19 patients were recruited (Age range: 18∼62; 17 males and 7 females). Patients were randomly divided into three groups (Gr): Gr. 1 control (n=8); Gr. 2: AFO-202 beta glucan (n=8); and Gr. 3, a combination of AFO-202 and N-163 beta glucans (n=8). All three groups received the standard care while groups 2 and 3 received additional supplementation of beta glucans for 30 days. In addition to basic clinical parameters, we periodically evaluated D-Dimer, IL-6, erythrocyte sedimentation rate (ESR), C-reactive protein (CRP), the neutrophil to lymphocyte ratio (NLR), the lymphocyte to CRP ratio (LCR) and the leukocyte-CRP ratio (LeCR).

**Results:** The duration of hospital stay for all three groups was nearly equivalent. There was no mortality of the subjects in any of the groups. Intermittent oxygen was administered from day of admission for up to four to five days with mask (two to four Lpm) to two subjects in Gr. 2 and one subject in Gr. 3. None of the subjects required ventilation. The D-Dimer values in Gr. 1, which was on average 751 ng/ml at baseline, decreased to 143.89 ng/ml on day 15, but increased to 202.5 ng/ml on day 30, which in groups 2 and 3 decreased on day 15 and continued to remain at normal levels until day 30. IL-6 levels decreased on day 15 from an average of 7.395 pg/ml to 3.16 pg/ml in the control, 26.18 pg/ml to 6.94 pg/ml in Gr. 2 and 6.25 pg/ml to 5.22 pg/ml in Gr. 3. However, when measured on day 30, in Gr. 1, the IL-6 increased to 55.37 pg/ml while there was only slight marginal increase in Gr. 2 but within normal range, and the levels further decreased to less than 0.5 pg/ml in Gr. 3. The same trend was observed with ESR. LCR and LeCR increased significantly in Gr. 3. NLR decreased significantly in groups 2 and 3. There was no difference in CRP within the groups.

**Conclusion:** In this exploratory study, consumption of Aureobasidium pullulans produced beta glucans for thirty days, results in a significant control of IL6, D-Dimer and NLR, a significant increase in LCR, LeCR and marginal control of ESR in COVID-19 patients. As these beta glucans are well known food supplements with decades of a track record for safety, based on these results, we recommend larger multi-centric clinical studies to validate their use as an adjunct in the management of COVID-19 and the ensuing long COVID-19 syndrome.

## Introduction

COVID-19 is caused by the novel strain of enveloped RNA virus, SARS-CoV-2 [1] with symptoms including fever, cough, shortness of breath, fatigue, body aches, headache, loss of taste or smell, sore throat, congestion or runny nose, nausea or vomiting and diarrhoea. The COVID-19 is an ongoing pandemic with nearly 200 million people affected worldwide and more than four million deaths to date [2]. Many patients are hospitalized 10 to 12 days after a positive RT-PCR test [3]. Recovery occurs within 14 days in most of the affected patients, but it may take more than 25 days in some patients [4]. Even patients with mild symptoms may progress to requiring oxygen therapy and mechanical ventilation and/or leading to death. The progression to severe disease is attributed to the hyperactive inflammatory response leading to a cytokine storm, thereby causing organ damage [5]. A dysfunctional immune-coagulation response activating the coagulation cascade leading to a hypercoagulable state may cause adverse clinical outcomes including death. Patients with co-morbidities such as diabetes, dyslipidemia and risk of coagulation are prone to a rapid progression of the disease and an immune-inflammatory-coagulation dysregulation-related adverse outcome [5]. Several interventions are part of the standard treatment regimen, including pharmacological agents such as anti-virals, IL6 inhibitors, anti-coagulants, steroids and supplements such as vitamins or zinc [6]. The solidarity trial, an international clinical trial to help find an effective treatment for COVID-19 continues to generate evidence for the use of these interventions, and the search for effective COVID-19 therapeutics continues [6]. Many of the pharmacological interventions have associated side effects, and in regard to the supplements, zinc or ascorbic acid have not been able to significantly decrease the duration of symptoms compared with standard care [3].

Given this background, the biological-response modifier glucans (BRMG) such as the *Aureobasidium pullulans* produced β-glucans have been suggested as an alternative adjunct treatment based on their potential to activate both the innate and adaptive arms of the host immune response against COVID-19 [7, 8]. This BRMG has been reported to reduce the levels of IL-1β, IL-2, IL-4, IL-6, IL-12, TNF-α, IFN-γ, and sFasL while increasing IL8 and sFAS, thereby exerting an effective optimal defence against viral infection without hyperinflammation [9]. The metabolic effects of the A. pullulans beta glucan in normalizing blood glucose and lipid levels add to their benefits for use in COVID-19 as fasting blood glucose is an independent outcome of COVID-19 severity and mortality [10]. The potential of this BRMG in acting as a prophylactic supplement to help combat coagulopathy associated with COVID-19 has also been described [11].

In an animal study using healthy SD rats, AFO-202 beta glucan was beneficial in decreasing neutrophil to lymphocyte ratio (NLR) while increasing lymphocyte to CRP ratio (LCR) and the leukocyte-CRP ratio (LeCR) in 15 days [12], whereas in a non-alcoholic steatohepatitis (NASH) disease animal model, it has been demonstrated that N-163 beta glucan has the potential for anti-inflammatory, anti-fibrotic immune modulation [13]. Similarly, in healthy human volunteers, the reported decrease in pro-inflammatory markers and increase in anti-inflammatory markers in an advantageous manner with the same beta glucans [14], thus supporting their ability to aid recovery in COVID-19 was considered worth studying. In this report, we hereby present the evaluation of the beneficial outcome of β-glucans produced by *A. pullulans* (AFO-202 and N-163 strains) in patients with COVID-19.

## Methods

The present study was an open label, prospective, randomized, comparative, multiple arm pilot clinical study to evaluate the immune enhancement and immunomodulatory efficacy of supplementation with *A. pullulans’* novel strains produced beta glucans compared to those who undergo a conventional therapeutic regimen alone in adult subjects with COVID-19 caused by SARS-CoV2(B-CoV).

Adult subjects between 18 and 65 years (both ages and sexes inclusive) who were confirmed to be positive for SARS-CoV2 by way of RT-PCR with or without co-morbidities, and with mild to moderate COVID-19 symptoms [15] but who required hospitalization were included in the study. Severe COVID-19-affected patients requiring intensive care, children and pregnant women were excluded.

There were three groups (Gr.) comprised of eight subjects each (Total n =24).

**Gr. 1 (n=8):** The control group receiving the standard treatment, which consisted of Inj. Remdesivir 200 mg (Day 1), Inj. Remdesivir 100 mg (Day 2 to Day 5) Inj. Solumedrol 80mg IV BD Inj. Clexane 40mg OD, broad spectrum antibiotic bronchodilators and supportive measures.

**Gr. 2 (n=8):** The intervention was standard treatment along with AFO-202 beta glucan supplement at 3 gm per day (1.0 gm granule in a sachet containing 42mg active ingredient of ß-1,3-1,6 Glucan, with each meal).

**Gr. 3 (n=8):** The intervention was AFO-202 beta glucan at 3 gm per day (1.0 gm granule with each meal) in combination with N-163 beta glucan at 10 gm per day (10 gm gel in a sachet with 90mg of ß-1,3-1,6 Glucan, with one of the meals every day).

The primary outcome was improvement in the clinical symptoms of COVID-19: time taken for improvement, complete recovery, recurrence in typical symptoms from baseline.

The secondary outcome was evaluation for required hospitalization, mortality, progression to critical care admission, oxygen/life-support and tests for biochemical parameters such as D-Dimer, IL6, ESR, CRP, NLR, LCR and LeCR.

Data were analysed using Microsoft Excel and Origin 2021b statistical software. Results are presented as mean or mean ± standard deviation for the continuous normal variables. Independent sample T-test and Kruskal-Wallis Test was used for comparison between the groups. A p-value < 0.05 was considered significant.

## Results

The baseline characteristics of the patients are presented in Table 1. The age of the subjects in Gr. 1 Control was 33 to 59 years (Mean = 47.62 years); in Gr. 2, AFO-202, 18 to 50 years (Mean = 36.25 years); and in Gr. 3, AFO 202+N163, 26 to 60 years (Mean= 39.87 years). There were 17 males and seven females recruited, and therefore the distribution was not equal across the groups. There were no significant differences among the groups in terms of body weight, BMI and blood pressure at the time of admission. All enrolled patients had mild to moderate COVID-19 [15] and none required ventilation or ICU admission. The average duration of stay in the hospital was 4.25 days in Gr. 1, 4.75 days in Gr. 2 and 4.125 days in Gr. 3. Two to four litres of oxygen were administered in two patients in Gr. 2 and in one patient in Gr. 1 from admission up to four or five days during the hospital stay. There was no statistically significant difference among the groups in terms of duration of hospitalization or the resolution of symptoms. There was no mortality in any of the groups. There was one serious adverse effect (SAE) in Gr. 1 (Control). The subject’s ECG on Visit 2 showed abnormalities after which an angiogram was taken which showed spontaneous coronary artery dissection in the left anterior descending portion of the right coronary artery. The subject was managed as per standard care under the supervision of a cardiologist.

**Table 1:** Baseline characteristics of study subjects

| Groups | Subject | Age range (years) | Gender | Body weight (Kgs) | Height (Cm) | BMI | Duration of hospital stay until recovery (No. of days) | BP (mm/Hg) |
| --- | --- | --- | --- | --- | --- | --- | --- | --- |
| Gr.1 - Control | 1 | 51-55 | Male | 80 | 175 | 26.1 | 3 | 130/80 |
|  | 2 | 41-45 | Female | 65 | 162 | 24.8 | 3 | 110/90 |
|  | 3 | 51-55 | Female | 75 | 170 | 26 | 5 | 100/70 |
|  | 4 | 51-55 | Female | 62 | 170 | 21.5 | 6 | 110/ 70 |
|  | 5 | 51-55 | Male | 78 | 175 | 25.5 | 4 | 130/90 |
|  | 6 | 46-50 | Female | 78 | 168 | 27.6 | 4 | 120/90 |
|  | 7 | 36-40 | Female | 70 | 152 | 30.3 | 5 | 120/80 |
|  | 8 | 31-35 | Female | 53 | 152 | 22.9 | 4 | 130/90 |
| Gr. 2- AFO-202 | 9 | 26-30 | Male | 60 | 156 | 24.7 | 6 | 120/80 |
|  | 10 | 26-30 | Male | 82 | 167 | 29.4 | 6 | 110/90 |
|  | 11 | 16-20 | Male | 81 | 180 | 25 | 6 | 110/90 |
|  | 12 | 46-50 | Male | 71 | 156 | 29.2 | 5 | 130/80 |
|  | 13 | 46-50 | Male | 80 | 170 | 27.7 | 3 | 120/90 |
|  | 14 | 41-45 | Male | 75 | 168 | 26.6 | 2 | 110/70 |
|  | 15 | 46-50 | Male | 68 | 168 | 24.1 | 4 | 120/90 |
|  | 16 | 31-35 | Male | 56 | 158 | 22.4 | 6 | 130/90 |
| Gr. 3- AFO-202 + N-163 | 17 | 26-30 | Male | 72 | 170 | 24.9 | 4 | 110/90 |
|  | 18 | 31-35 | Male | 55 | 150 | 24.4 | 5 | 120/90 |
|  | 19 | 61-65 | Male | 75 | 176 | 24.2 | 4 | 110/70 |
|  | 20 | 26-30 | Male | 80 | 172 | 27 | 3 | 130/90 |
|  | 21 | 36-40 | Male | 76 | 174 | 25.1 | 3 | 130/90 |
|  | 22 | 26-30 | Male | 79 | 174 | 26.1 | 3 | 120/90 |
|  | 23 | 41-45 | Female | 79 | 160 | 30.9 | 2 | 120/90 |
|  | 24 | 51-55 | Male | 84 | 166 | 30.5 | 9 | 110/70 |

The drop-outs were high in the 30-day follow-up (11 subjects lost to follow up: three in Gr. 1 and four in groups 2 and 3 each) (figure 1) due to a reluctance in the subjects to come to the hospital for follow-up as most of them were worried about re-infection and psychological factors on returning to the hospital from where they were discharged. The oxygen saturation increased in all the groups in the 15- and 30-day follow-up but the difference was not significant. The temperature and heart rate also showed no significant difference among the groups (figure 2).

**Figure 1:**
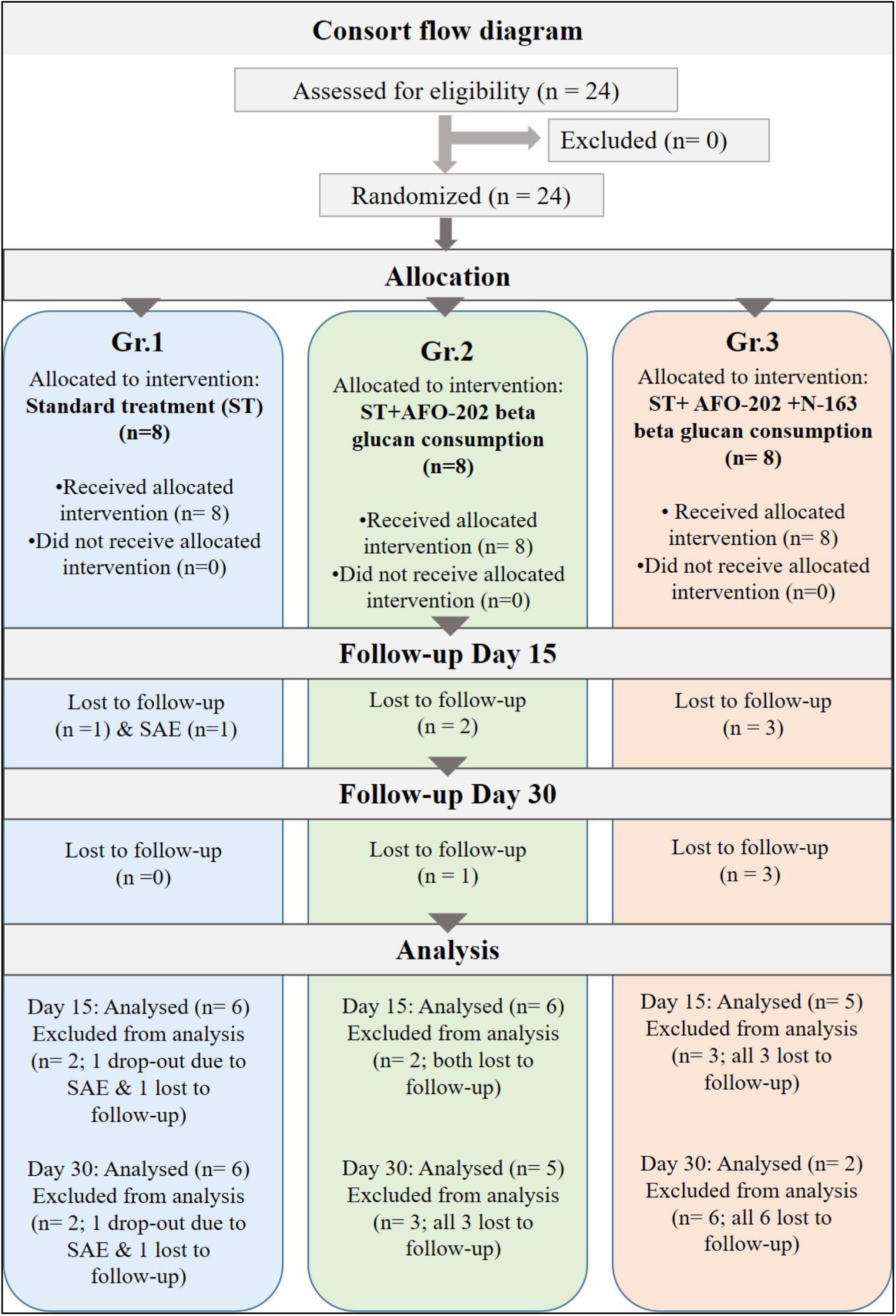
CONSORT flow diagram of the trial.

**Figure 2:**
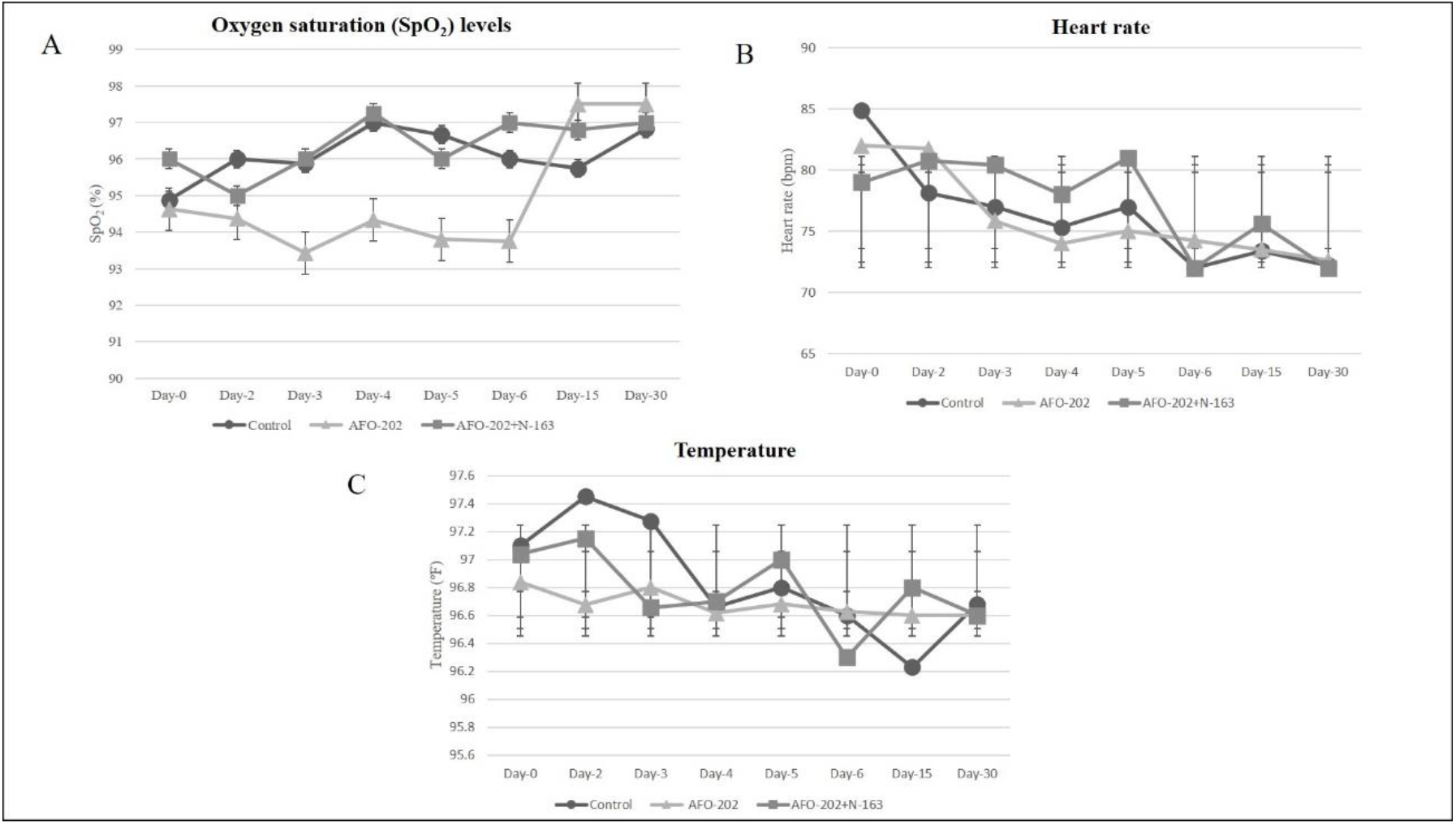
A. Oxygen saturation (SpO2) (%) B. Heart rate (bpm) and C. Temperature (ºF) and at baseline (Day 0), day 2, day 3, day-4, day-5, day-6, day 15 and day 30 in the different treatment arms of COVID-19 subjects.

In the control group, the mean fasting blood glucose (FBG) levels on day 0 were 210.14 mg/dl which decreased to 121.5 mg/dl on day 30. In Gr. 2, the mean FBG decreased from 154.14 mg/dl to 103 mg/dl in 30 days. In Gr. 3, it decreased from 238 mg/dl to 140 mg/dl in 30 days (figure 3). The difference among the groups was not significant.

**Figure 3:**
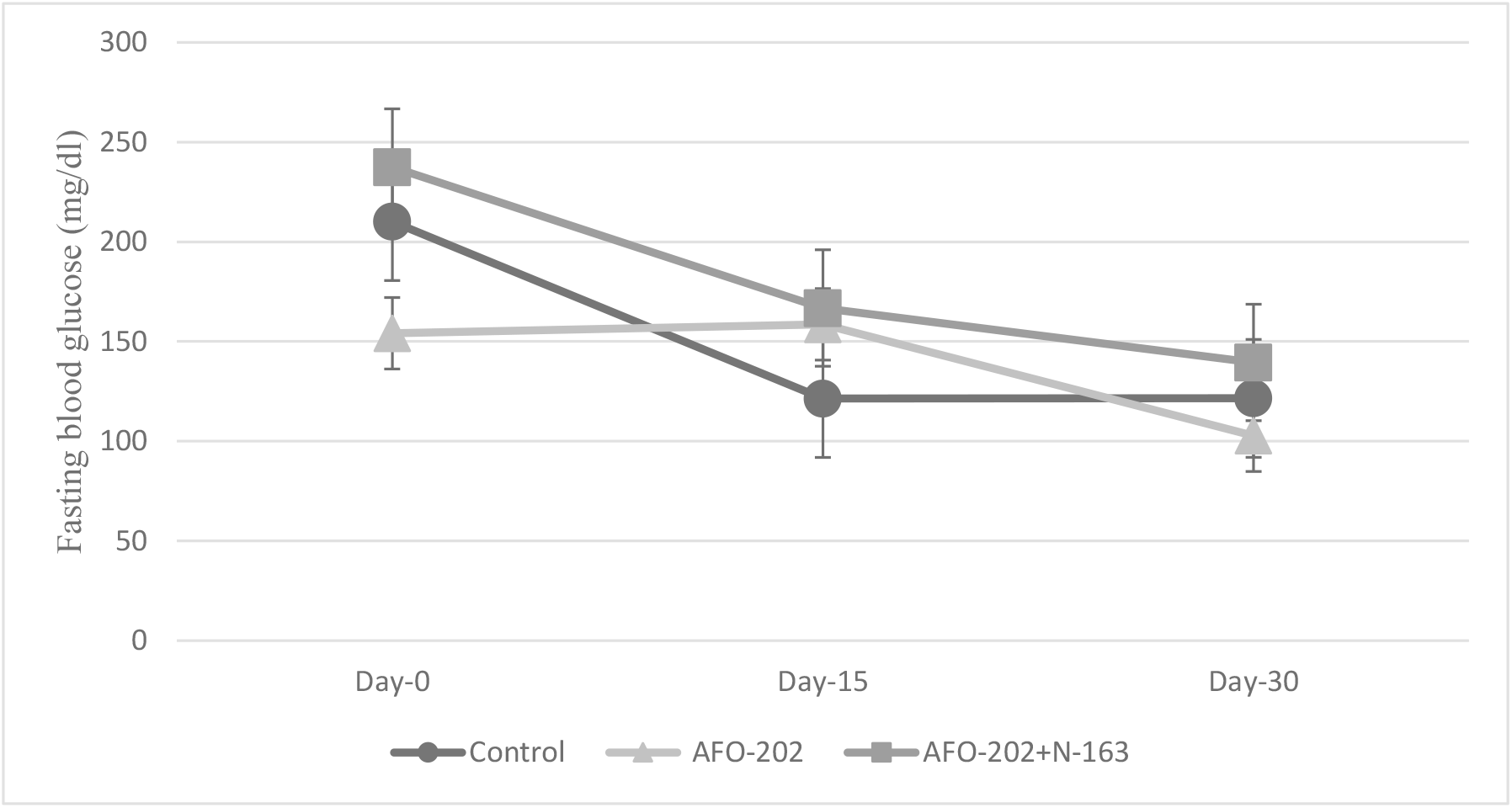
Fasting blood glucose levels at baseline (Day 0), day 15 and day 30 in the different treatment arms of COVID-19 subjects showing decrease in the all groups.

In the control group, IL6 values decreased from an average of 7.395 pg/ml to 3.16 pg/ml in day 15 but again increased to 55.37 pg/ml at day 30. However, in Gr. 2 (AFO-202), IL6 levels steadily decreased from a mean of 26.18 pg/ml to 6.94 pg/ml in 15 days and to 3.41 pg/ml in 30 days, while in Gr. 3 (AFO202+N163) it decreased from 6.25 pg/ml to 5.22 pg/ml in 15 days and to 0.5 pg/ml in 30 days. The results were statistically significant (p value =0.0214) (figure 4).

**Figure 4:**
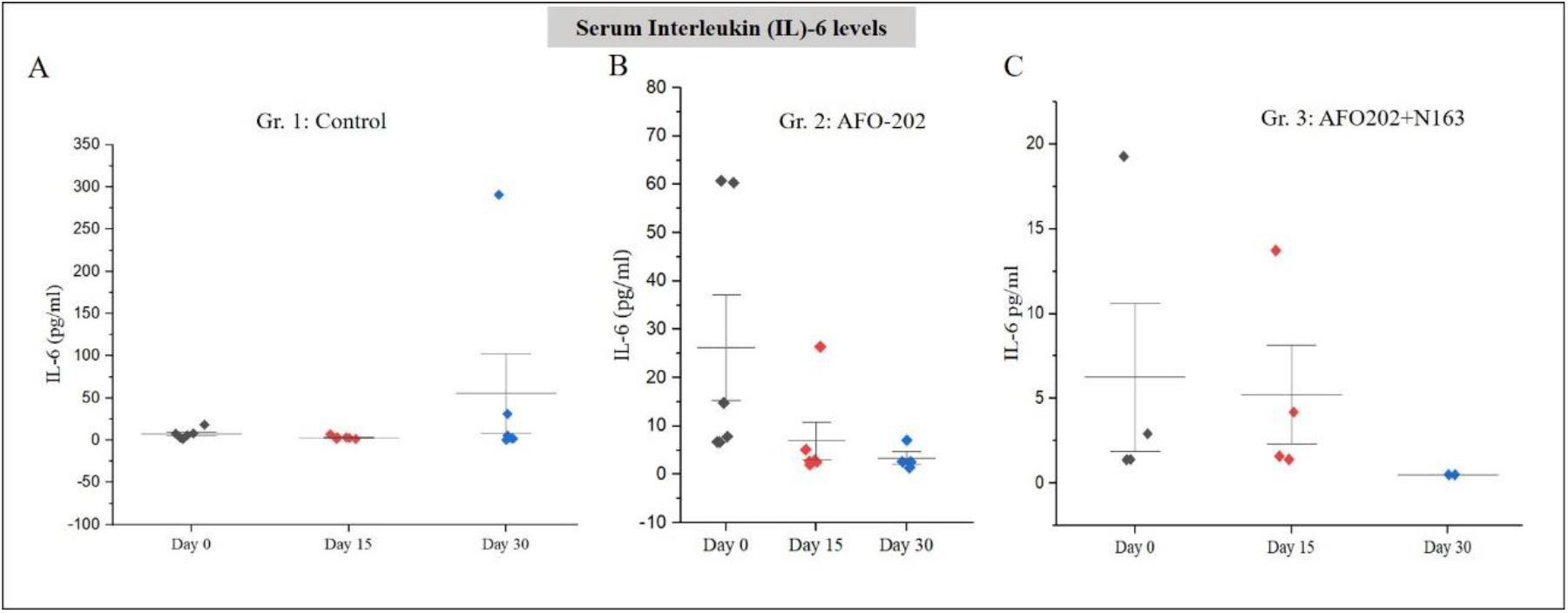
Serum Interleukin (IL)-6 levels in subjects with COVID-19 A. Gr. 1: Control (Standard treatment); B. Gr. 2: (Standard treatment + AFO-202 beta glucan supplementation); and C. Gr. 3: (Standard treatment + AFO-202 and N-163 beta glucan supplementation) at baseline (day 1), day 15 and day 30 wherein the decrease was statistically significant in Gr.2 and 3 (p < 0.05).

In Gr. 1 patients, levels of D-Dimer decreased until day 15 but then increased on day 30 (mean = 751 ng/dl to 143.89 ng/dl on day 15 and 202.5 ng/dl on day 30). In contrast, in groups 2 and 3, the levels of D-Dimer steadily decreased until day 30. In Gr. 2 on Day 0, IL6 was 560.99 ng/dl, while on Day 30, it was 79.615 ng/dl. In Gr. 3 on day 0, it was 1614 ng/dl to 164.25 ng/dl on day 30. This was statistically significant (p value = 0.013) (figure 5).

**Figure 5:**
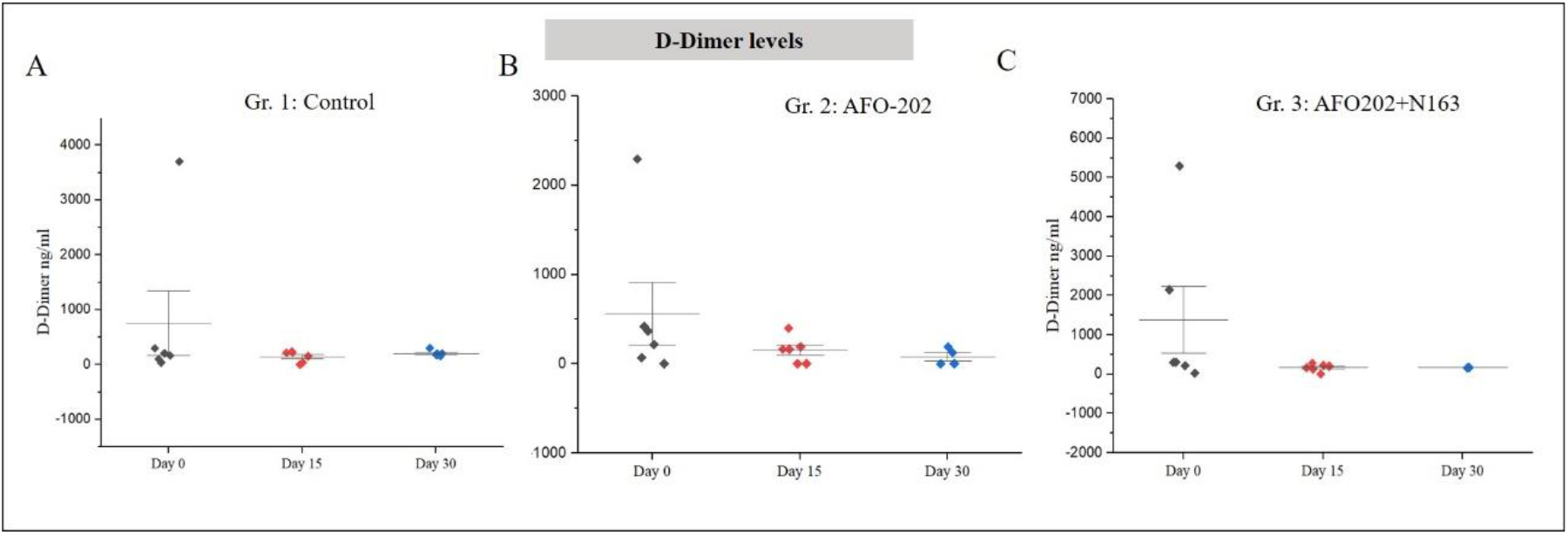
D-Dimer Levels in subjects with COVID-19 A. Gr. 1: Control (Standard treatment); B. Gr. 2: (Standard treatment + AFO-202 beta glucan supplementation); and C. Gr. 3: (Standard treatment + AFO-202 and N-163 beta glucan supplementation) at baseline (day 0), day 15 and day 30 wherein the decrease was statistically significant in Gr.2 and 3 (p < 0.05).

While CRP showed a steady decrease from day 0 to day 15 and day 30 in all the groups (figure 6), ESR in Gr. 1 decreased when measured on day 15 but increased on day 30 in Gr. 1. In Gr. 2 and 3 the ESR values were lower than in control patients on day 15 and day 30, although this was not statistically significant (figure 7).

**Figure 6:**
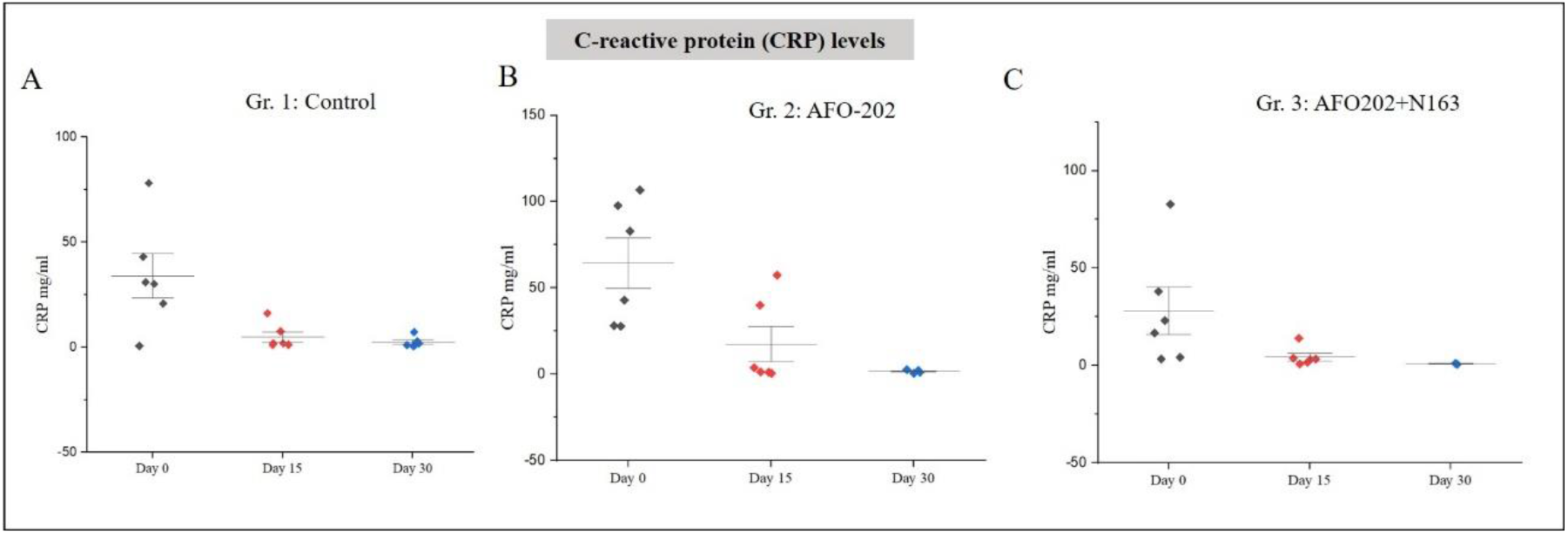
C-reactive protein (CRP) Levels in subjects with COVID-19 A. Gr. 1: Control (Standard treatment); B. Gr. 2: (Standard treatment + AFO-202 beta glucan supplementation) and C. Gr. 3: (Standard treatment + AFO-202 and N-163 beta glucan supplementation) at baseline (day 0), day 15 and day 30.

**Figure 7:**
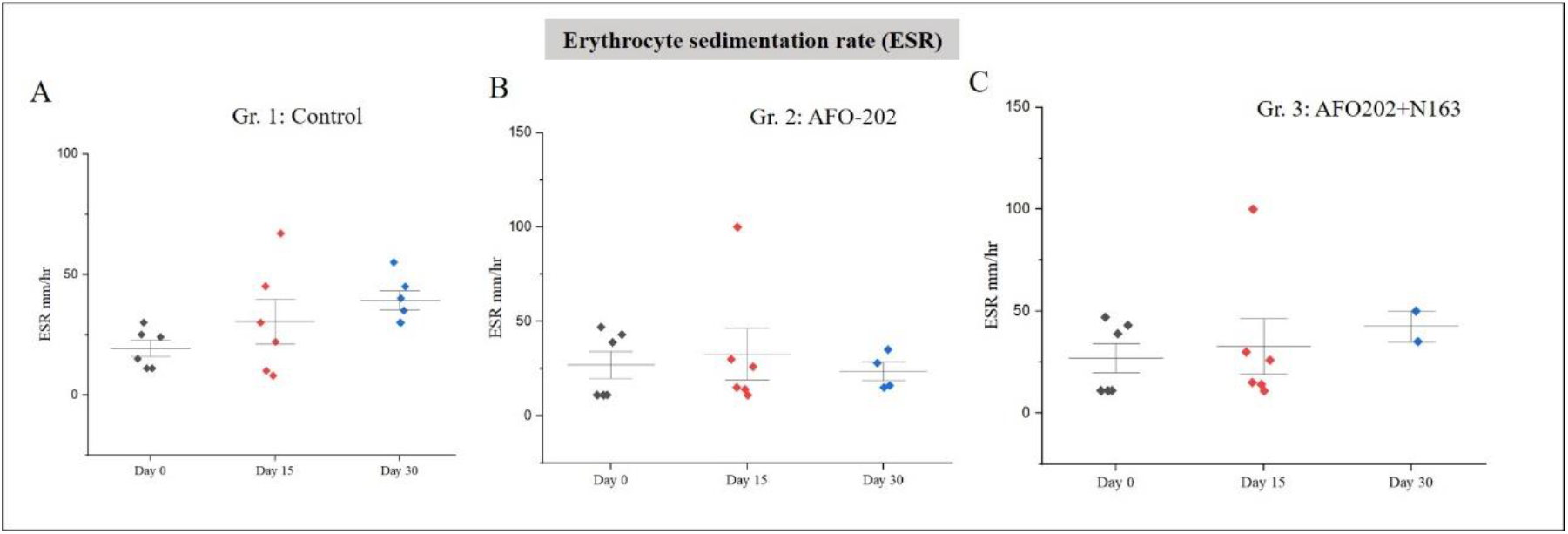
Erythrocyte sedimentation rate (ESR) values in subjects with COVID-19 A. Gr. 1: Control (Standard treatment), B. Gr. 2: (Standard treatment + AFO-202 beta glucan supplementation); and C. Gr. 3: (Standard treatment + AFO-202 and N-163 beta glucan supplementation) at baseline (day 0), day 15 and day 30.

The decrease in NLR (figure 8) and the increase in LCR and LeCR (figure 9) was greater in Gr. 3 at both 15 days and 30 days compared to the other groups.

**Figure 8:**
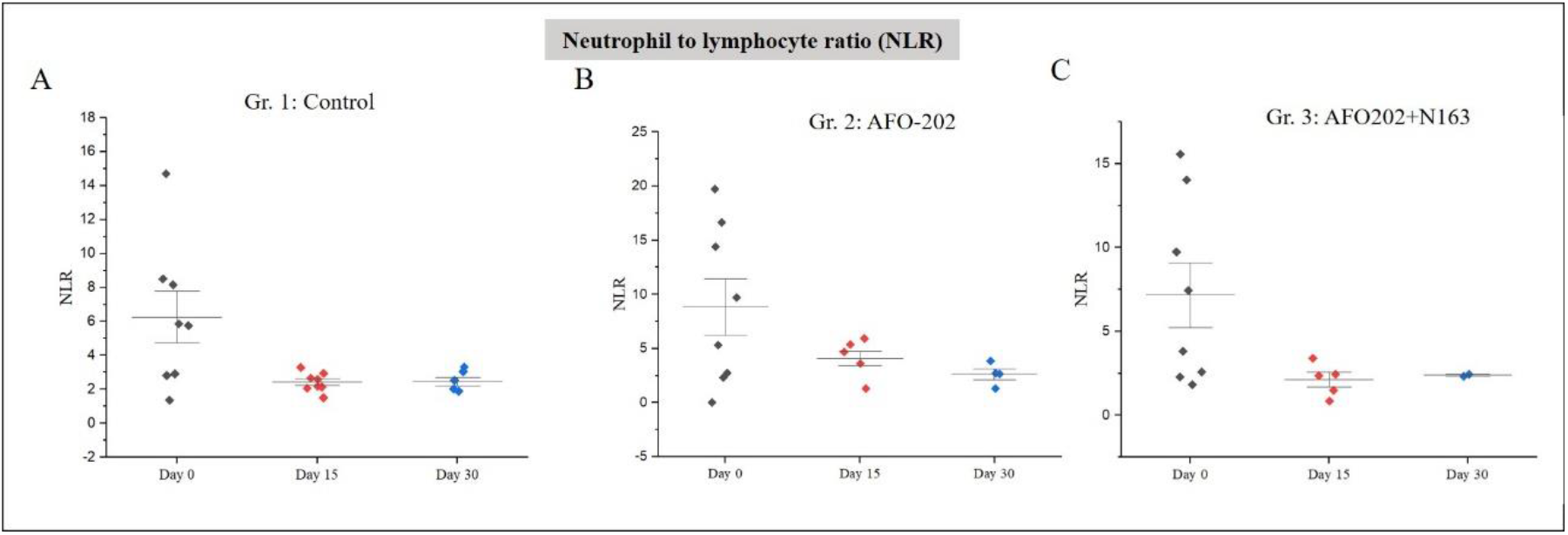
Decrease in neutrophil to lymphocyte ratio (NLR) values in subjects with COVID-19 A. Gr. 1: Control (Standard treatment), B. Gr. 2: (Standard treatment + AFO-202 beta glucan supplementation) and C. Gr. 3: (Standard treatment + AFO-202 and N-163 beta glucan supplementation) at baseline (day 0), day 15 and day 30.

**Figure 9:**
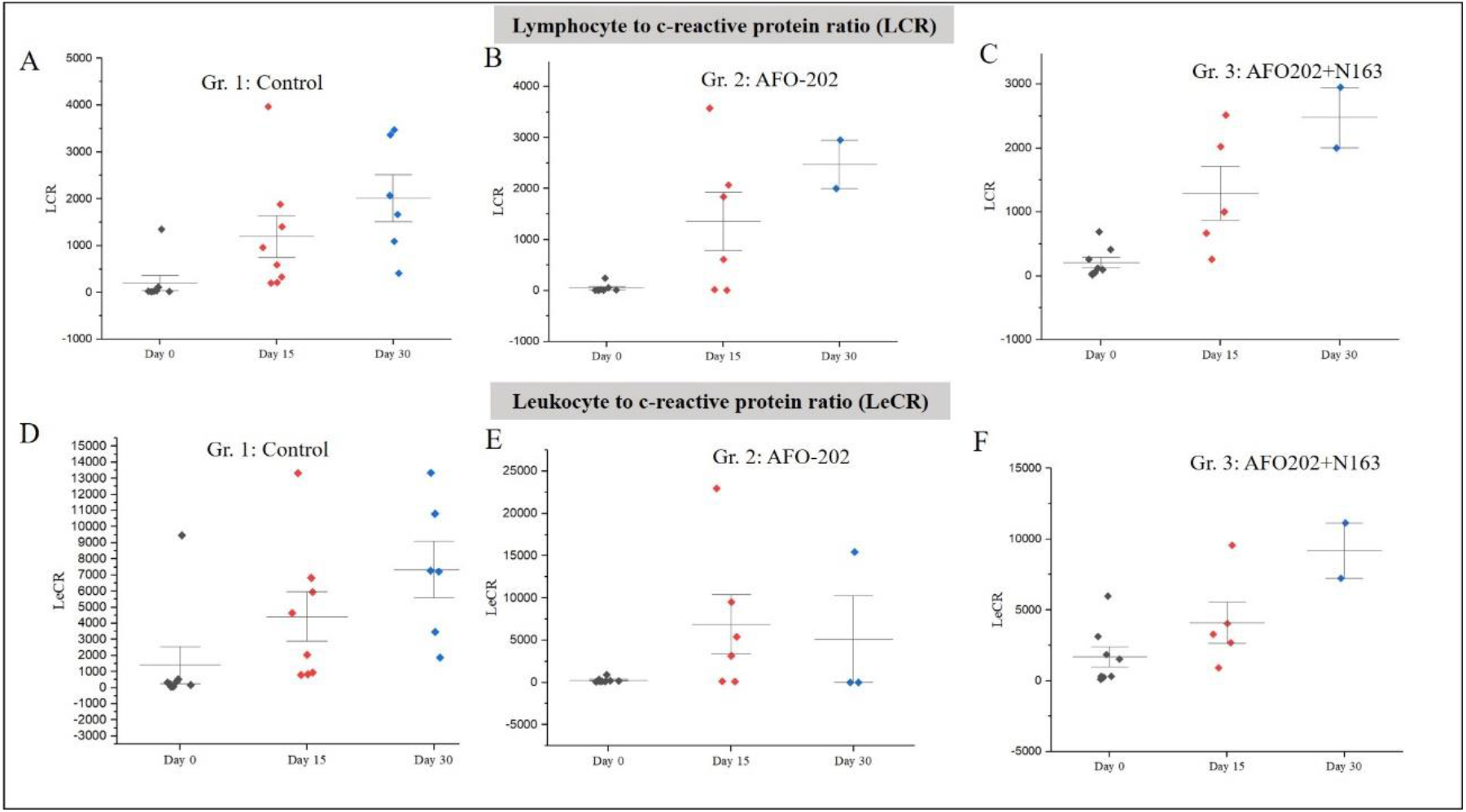
Increase in A-C: Lymphocyte to c-reactive protein (LCR) values in subjects with COVID-19 in A. Gr. 1: Control (Standard treatment); B. Gr. 2: (Standard treatment + AFO-202 beta glucan supplementation); C. Gr. 3: (Standard treatment + AFO-202 and N-163 beta glucan supplementation); D-F: Leukocyte to c-reactive protein (LeCR) values in subjects with COVID-19 D. Gr. 1: Control (Standard treatment); E. Gr. 2: (Standard treatment + AFO-202 beta glucan supplementation); and F. Gr. 3: (Standard treatment + AFO-202 and N-163 beta glucan supplementation) at baseline (day 0), day 15 and day 30.

## Discussion

The immunological profile from pathogenesis to progression to a severe disease in COVID-19 leading to a cytokine storm assessed by elevation of biomarkers such as IL6 and coagulopathy assessed by the elevation of markers such as D-Dimer has been reported to be unique [16]. Though many recover after one or two weeks of illness, the second to fourth week after initial onset has been reported to be stormy as many develop immune hyper-activation leading to cytokine storm marked by elevation of markers such as NLR and CRP, decrease of LCR and LeCR [17], or coagulopathy disruption evident by elevation of markers such as D-Dimer and ferritin. Standard care of management that has been recommended by professional bodies includes steroid based immune suppression or anti-coagulants [6]. Even under appropriate treatment, morbidity, mortality and related correlations to the pathogenesis remain uncertain [6]. There have been reports of bleeding diathesis after anti-coagulants [18], and the appropriate timing for administration of these pharmacological agents are still being debated [19]. At this hour, a safer and more efficient interventional strategy to modulate the immune system to prevent or tackle the cytokine storm and to manage the coagulopathy cascade is needed.

A balanced immune enhancement with metabolism regulation potential, of AFO-202 beta glucan has been reported earlier, while an efficient immune-modulation including anti-fibrotic effects has been reported using N-163 beta glucan [7–14]. Specially, in healthy volunteers’ alleviation of glucotoxicity by balancing of blood glucose with AFO-202 and alleviation of lipotoxicity by balancing of lipid profile when AFO-202 beta glucan is combined with N-163 beta glucan has been reported [14]. Such combinations yielding beneficial effects in animal studies [13] of NASH models have proven the anti-fibrotic and anti-inflammatory immunomodulatory capabilities. The above studies in healthy animal models, human volunteers and diseased animal models have demonstrated the potentials of these beta glucans offering a balanced immunomodulation along with the necessary immune enhancement. Having used them alone or in combination, we report results herein in COVID-19 patients yielding both benefits, such as balanced modulation of inflammatory associated parameters including IL-6, NLR, LCR and LeCR. However, in addition to the beneficial effects on the immune system, the added benefits in regulating the coagulopathy evident by decrease in D-Dimer is a unique dual advantage which has never been reported with a single standalone agent or a molecule for COVID-19 in the literature, to our knowledge.

D-dimer has been reported to have the highest C-index to predict in-hospital mortality in COVID-19 patients [5]. A D-dimer level greater than 0.5 μg/ml is associated with severe infection, and a value greater than 1 μg/ml is associated with increasing odds of in-hospital death in patients with COVID-19 [20]. In the current study, the combination of AFO-202 and N-163 beta glucans were able to bring down the D-Dimer values which was greater than 1.6 μg/ml at baseline to normal values in 15 days (figure 5) that continued to be maintained even until day 30 against the control group in whom it increased. Though all the participants received anti-coagulants, the normalization of D-Dimer was significant when supplemented with beta glucans as in Gr.2 and 3.

Likewise, IL-6 levels have been reported to be significantly elevated and associated with adverse clinical outcomes in COVID-19. Inhibition of IL-6 has been suggested as a novel target for managing dysregulated host responses in patients with COVID-19 [21]. In the current study, the IL-6 values were decreased from values predictive of higher mortality due to COVID-19 > 24 pg/ml [22] to normal values which were maintained in the beta glucan groups only while it increased to values predictive of higher mortality in the control groups at day 30 (figure 4). The significant and steady decrease in IL6 and D-Dimer (figure 4 and 5) substantiate the anti-inflammatory and anti-coagulation benefits of beta glucans. The observation that patients in groups 2 and 3 had normal levels of IL-6, D-Dimer and ESR in contrast to increased levels on day 30 in the control group strongly suggests their relevance to potential management of post-COVID or long-COVID manifestations [23]. Other significant predictors of severity in COVID-19 such as NLR, LCR and LeCR were also maintained at normal range in the beta glucan groups.

Although several other food supplements including n-3 PUFA, zinc, and vitamins C and D, are being investigated for their potential applications in COVID-19 [3], the clinical trial results have largely yielded no positive outcome. Probiotics have better advantages than the other dietary supplements because of their effects at the entry point of the SARS-CoV2 virus in the intestinal epithelium and the lung-gut axis [24–26]. Beta glucans are beneficial in terms of the pre-biotic effects on the gut microbiota as well [27] which could play a significant role contributing to mechanisms by which they exert their influence on the mucosal and other arms of immunity including the trained immune response [28], which needs additional elaborate research.

The safety profile of both AFO-202 and N-163 beta glucans has been further corroborated by the no SAEs or mortality in groups 2 and 3. The increase in the biomarkers for inflammation (IL6, NLR, ESR, D-Dimer) in spite of standard treatment and low molecular weight heparin administration in Gr. 1 but not in groups 2 and 3 confirms the beneficial effects of the beta-glucans as advantageous adjuncts to presently available standard treatments of COVID-19. The metabolic benefits with immune enhancement of AFO-202 alone, which when combined with N-163 beta glucans yielding a stronger immunomodulation and anti-inflammatory effect has been significantly demonstrated in the current study.

The limitations of the study include the large number of drop-outs that have been attributed to the psychosocial factors of visiting the hospital for follow-up. In addition, the sample number was less, but this is only a pilot clinical study. We conducted this study at the time of a decline in the second wave in India [29] when infection due to the delta variant of SARS-CoV2 virus was gradually emerging as a threat [30] with increased contagiousness [31]. We have not studied genomic sequencing in the subjects to identify the delta variant virus, and this needs further evaluation to study the efficacy of the beta glucans against emerging variants. The significance of beta glucan groups versus control though is evident in this study, the difference in the quantum of beneficial effects between the AFO-202 (Gr.2) and that of Gr. 3 due to smaller sample numbers is not very clear, as reported in the earlier clinical study in terms of immunomodulation in healthy volunteers [14] and anti-inflammatory, anti-fibrotic effects in NASH animal models [13], warrant a larger study to confirm the same. In such larger multi-centric clinical trials, evaluations on other biochemical and immunological parameters along with radiological diagnostics can help decipher the various underlying mechanisms to further validate the advantageous effects of these safety-proven beta-glucans so that they could be recommended for continued supplemental prophylaxis as a vaccine adjuvant [8] and as an adjunct to the existing treatments for the management of COVID-19.

The continuous increase in ESR, CRP, IL6 and D-Dimer on day 30 in the control group may be indicative of the post-Covid sequalae or the long COVID syndrome [23]. Having documented that the levels of these biomarkers were maintained in the normal range in the beta glucan groups even until day 30, we suggest their application in the management of long-COVID after necessary validation, with a longer duration of follow-up.

## Conclusion

Supplementation with the beta glucans produced by the AFO-202 and N-163 strains of Aureobasidium pullulans have helped to maintain the major biomarkers of clinical severity and mortality of COVID-19 viz, IL-6, D-Dimer, NLR over 15 and 30 days compared to those who underwent standard care alone in this randomized pilot clinical study. These findings add to the advantages of these nutraceutical agents as continuous oral supplemented adjuncts for prophylaxis and management of COVID-19, especially in those with co-morbid conditions. After necessary validation in larger multicentric studies, they have a potential for being recommended as an adjunct in the management of the long COVID syndrome or post-COVID sequalae.

## Data Availability

All data generated or analysed during this study are included in this manuscript.

## Acknowledgements

The authors wish to acknowledge,

a. The Government of Japan and the Prefectural Government of Yamanashi for a special loan for COVID-19 related implications and M/s Yamanashi Chuo Bank for processing the transactions.
b. The co-investigators Dr. Vaiyali Ramu, Emergency and Critical Care Physician, Shanmuga Hospital Pvt Ltd, India, Dr. S. R. Tiruvalavan, Clinical Research Head, Shanmuga Hospital Pvt Ltd, India, and their colleagues; Medical, nursing, para-medical, data collection - clerical staff for their valuable contribution to the clinical trial amidst the ongoing pandemic.
c. Mr. Takashi Onaka, Mr. Yasunori Ikeue and Dr. Mitsuru Nagataki (Sophy Inc, Kochi, Japan), for necessary technical clarifications.
d. Mr. Yoshio Morozumi and Ms. Yoshiko Amikura of GN Corporation, Japan for their liaison assistance with the conduct of the study.
e. Loyola-ICAM College of Engineering and Technology (LICET) for their support to our research work.

